# The effects of sleep disturbance on dyspnoea and impaired lung function following COVID-19 hospitalisation: a prospective multi-centre cohort study

**DOI:** 10.1101/2022.12.13.22283391

**Authors:** C. Jackson, I. Stewart, T. Plekhanova, P. Cunningham, A. L. Hazel, B. Al-Sheklly, R. Aul, C. E. Bolton, T. Chalder, J. D. Chalmers, N. Chaudhuri, A. B. Docherty, G. Donaldson, C. L. Edwardson, O. Elneima, N. J. Greening, N. A. Hanley, V. C. Harris, E. M. Harrison, L-P. Ho, L. Houchen-Wolloff, L. S. Howard, C. J. Jolley, M. G. Jones, O. C. Leavy, K. E. Lewis, N. I. Lone, M. Marks, H. J. C. McAuley, M. A. McNarry, B. Patel, K. Piper-Hanley, K. Poinasamy, B. Raman, M. Richardson, P. Rivera-Ortega, S. Rowland-Jones, A. V. Rowlands, R. M. Saunders, J. T. Scott, M. Sereno, A. Shah, A. Shikotra, A. Singapuri, S. C. Stanel, M. Thorpe, D. G. Wootton, T. Yates, R. G. Jenkins, S. Singh, W. D-C. Man, C. E. Brightling, L. V. Wain, J. C. Porter, A. A. R. Thompson, A. Horsley, P. L. Molyneaux, R. A. Evans, S. E. Jones, M. K. Rutter, J. F. Blaikley, PHOSP-COVID Study Collaborative Group

## Abstract

**Background:** Sleep disturbance is common following hospitalisation both for COVID-19 and other causes. The clinical associations are poorly understood, despite it altering pathophysiology in other scenarios. We, therefore, investigated whether sleep disturbance is associated with dyspnoea along with relevant mediation pathways.

**Methods:** Sleep parameters were assessed in a prospective cohort of patients (n=2,468) hospitalised for COVID-19 in the United Kingdom in 39 centres using both subjective and device-based measures. Results were compared to a matched UK biobank cohort and associations were evaluated using multivariable linear regression.

**Findings:** 64% (456/714) of participants reported poor sleep quality; 56% felt their sleep quality had deteriorated for at least 1-year following hospitalisation. Compared to the matched cohort, both sleep regularity (44.5 vs 59.2, p<0.001) and sleep efficiency (85.4% vs 88.5%, p<0.001) were lower whilst sleep period duration was longer (8.25h vs 7.32h, p<0.001). Overall sleep quality (effect estimate 4.2 (3.0–5.5)), deterioration in sleep quality following hospitalisation (effect estimate 3.2 (2.0–4.5)), and sleep regularity (effect estimate 5.9 (3.7–8.1)) were associated with both dyspnoea and impaired lung function (FEV^1^ and FVC). Depending on the sleep metric, anxiety mediated 13–42% of the effect of sleep disturbance on dyspnoea and muscle weakness mediated 29-43% of this effect.

**Interpretation:** Sleep disturbance is associated with dyspnoea, anxiety and muscle weakness following COVID-19 hospitalisation. It could have similar effects for other causes of hospitalisation where sleep disturbance is prevalent.

**Funding:** UK Research and Innovation, National Institute for Health Research, and Engineering and Physical Sciences Research Council.

## Introduction

Persistent health changes following hospitalisation for COVID-19 are commonly reported and recognised as a post-COVID-19 syndrome^1^. For instance, 71% of participants in the post-hospitalisation COVID-19 study (PHOSP-COVID) reported persistent symptoms a median of 5 months following discharge^2^. In this setting, dyspnoea is a frequently reported symptom, occurring in 48% of cases^2^. Identifying the exact cause of dyspnoea post-hospitalization can be challenging as it can result from dysfunction in multiple organ systems e.g., neurological, respiratory, cardiovascular, and mental health^3^. Sleep disturbance also affects these organ systems^4^ and has been commonly reported following COVID-19 in some studies^5-14^. Despite this overlap, the association between sleep disturbance with dyspnoea has not been widely studied.

Sleep disturbance following hospitalization is common^15^ regardless of the original reason for admission. Despite its prevalence, the clinical implications of this condition following hospitalisation are still being described. The effects of general sleep disturbance, however, have been investigated with experimental models suggesting a causal association with both anxiety^16,17^ and muscle weakness^18^. Furthermore, clinical studies have revealed that general sleep disturbance is associated with chronic respiratory diseasese^19-21^. Whether these associations are still observed when sleep disturbance occurs acutely following hospitalisation has still to be fully explored.

Using one method to assess sleep disturbance is not ideal as each method has important limitations. Subjective assessments provide an overall score of sleep quality but can be affected by recall bias^22^ and questionnaire language. Subjective assessments also provide only limited insights into specific types of sleep disturbance. In contrast, device-based assessments of sleep quality e.g., actigraphy^23^, measure sleep disturbance subtypes but they do not assess overall sleep quality^24,25^. Therefore combining both subjective and device-based measures into a multi-modal approach^26^ can provide valuable insights into sleep disruption partially overcoming the limitation of individual approaches.

A limited number of studies^5-14^ have reported altered sleep quality following COVID-19 hospitalisation. Although some have been multi-centre, the majority are often single-centre, modest in size and used subjective measures. Two studies to date have employed a multi-modal approach^27,28^ to estimate sleep disturbance. In these studies, an association with anxiety was reported only with subjective but not device-based measures. Furthermore, no other clinical associations were reported. Finally, these studies only used participants that had been admitted to critical care, limiting generalisation to the broader hospital cohort.

Therefore, we aimed to characterise the prevalence, type, and relevance of sleep disturbance in a broad cohort of patients that had been hospitalised for COVID-19 using a multi-modal approach. We hypothesised that, in line with other studies, sleep disturbance would have a high prevalence. We also hypothesised, based on experimental models, that sleep disturbance would be associated with dyspnoea and that this association would be partially mediated by muscle weakness and anxiety.

## Methods

### Participants

All participants recruited into PHOSP-COVID were adults, ≥18 years of age, admitted to hospital with either PCR confirmed or clinically diagnosed COVID-19 and discharged between March 2020 – October 2021. The demographics and recruitment of participants into PHOSP-COVID have been described elsewhere^2^ and are briefly described in the supplementary methods. COVID-19 severity during admission was assessed using the WHO clinical progression scale^29^.

### Subjective assessment of sleep quality

This was assessed by two different methods a median of 5 months (IQR 4-6) post-hospitalisation.

i. **Pittsburgh Sleep Quality Index (PSQI) questionnaire**^30^: This questionnaire assesses sleep quality across seven components. A total score greater than 5 was defined as poor sleep quality and a score ≤ 5 was defined as good sleep quality^30^.
ii. **Numerical rating scale (NRS) assessment of sleep quality:** Patients were asked to rate their sleep quality on a numerical rating scale (0-10; zero being the worst sleep quality, **Suppl. Methods**).

### Device-based assessment of sleep quality

Participants were invited to wear a wrist-worn accelerometer (GENEActiv Original, ActivInsights, Kimbolton, UK) on their non-dominant wrist 24h/day for 14 days a median of 7 months (IQR 5-8) post-discharge. Sleep regularity, sleep efficiency, and sleep period duration were then analysed both continuously and split into quintiles with the top and bottom quintiles being used in subsequent analysis^27,28^. Further details of the data cleaning, analysis, and variable definitions are given in the supplementary methods.

### UK Biobank cohort

The UK Biobank^31^ was used as a pre-pandemic comparator cohort. The UK Biobank recruited 502,540 participants aged 40 – 69 years who were invited to a baseline visit at one of 22 assessment centres between 2006 and 2010 during which their phenotypes were established using questionnaires, physical examination, and collection of biological samples (**Suppl. Methods**).

### Symptom assessment

All symptoms were assessed at the early visit a median of 5 months (IQR 4 – 6) post-hospitalisation. Details of each assessment are given in the supplementary methods.

### Statistical analysis

Continuous values are presented as mean (95% CI) and ordinal values are presented as median (IQR). All univariable and multivariable analyses of continuous data were analysed using ordinary least squares linear regression. The multivariable analyses adjusted for a minimally sufficient set of covariates: age, sex, body mass index (BMI), comorbidities, COVID-19 severity, and length of stay. This set was identified based on a Directed Acyclic Graph (DAG; www.dagitty.net, **Suppl. Methods**). Multinomial logistic regression was used for modelling anxiety. The 95% confidence intervals for regression coefficients were calculated from a residual bootstrap approach with 1,999 resamples (**Suppl. Methods**). Chi-square tests compared the proportions of categorical variables. Mediation was evaluated using linear regression with the product of coefficients method^32^ (**Suppl. Methods**) to estimate the direct and indirect effects of the relationship, performed using the R package *lavaan* version 0.6-12. All data were analysed using R (version 4.2.0) within the Scottish National Safe Haven Trusted Research Environment. A *p*-value < 0.05 was considered significant.

## Ethics

The study was ethically approved (Ref: 20/YH/0225)

## Results

A total of 2,468 participants were enrolled in the PHOSP-COVID study and attended an early time point research visit, a median of 5 months (IQR 4-6) following discharge across 83 hospitals in the United Kingdom. Subjective sleep quality was measured using both the PSQI questionnaire and a numerical rating scale (NRS). 1,179 (51%) of participants attended an early follow-up at a centre offering the PSQI questionnaire (**Fig. 1**). Of these participants, 69% (812/1,179) completed the PSQI questionnaire and 88% (714/812) of PSQI respondents also completing the NRS at this early time point. At the late time point, 34% (277/812) of participants completed the NRS questionnaire (**Fig. 1**), a median of 12 months (IQR 11-13) after discharge. Device-based sleep quality was assessed using actigraphy in 38% (829/2,157) of participants (**Fig. 1**) a median of 7 months (IQR 5-8) after discharge.

**Figure 1:**
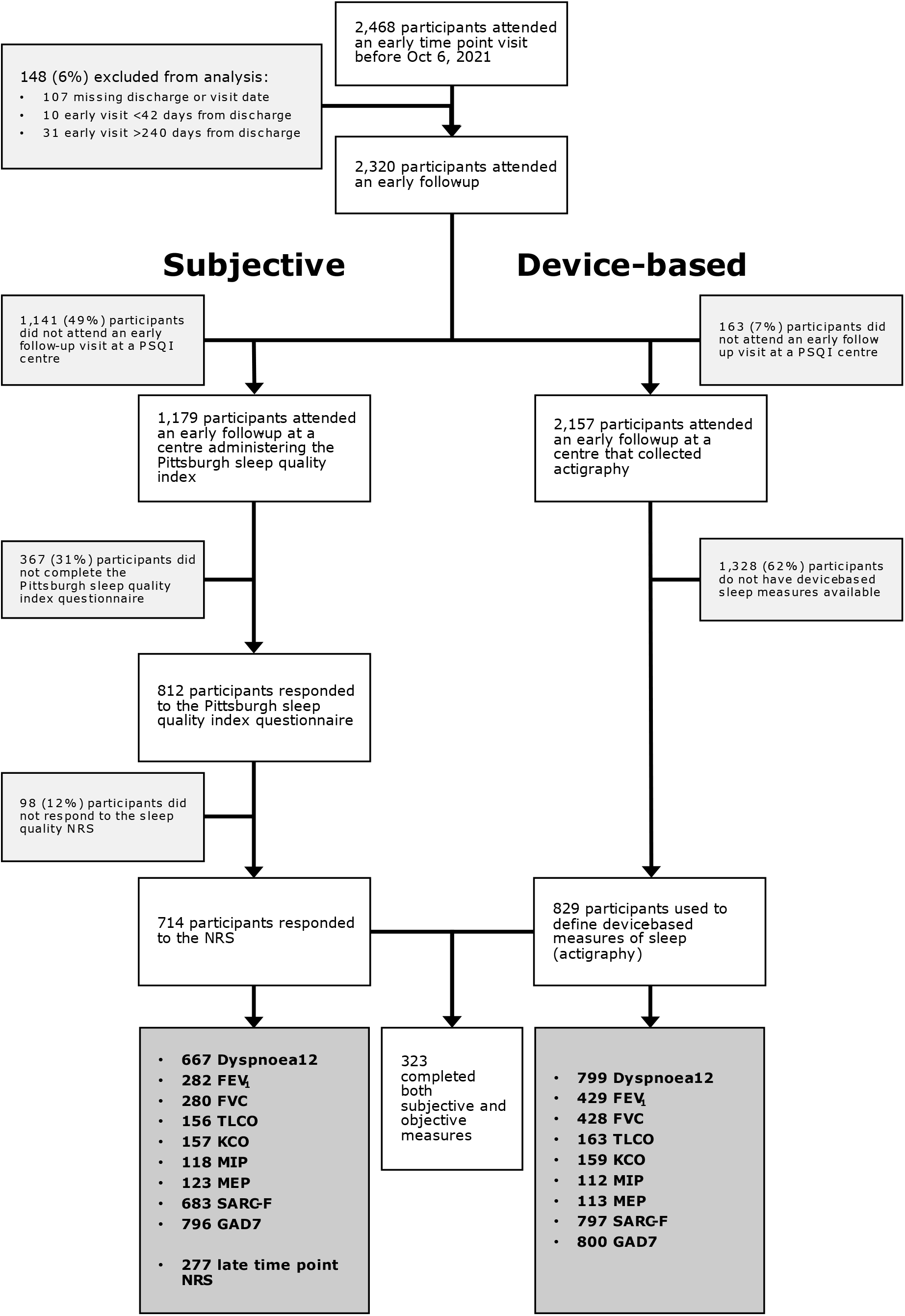
Consort diagram revealing the number of participants used in the analysis: Participants were recruited from the PHOSP-COVID study who were evaluated at the early time point and gave their consent for research. Sleep disturbance was evaluated using two types of measures (subjective and device-based). PSQI=Pittsburgh Sleep Quality Index. NRS=Numerical Rating Scale. FEV^1^=Forced Expiratory Volume in one second. FVC=Forced Vital Capacity. TLCO=gas transfer capacity. KCO=carbon monoxide transfer coefficient. MIP=Maximum Inspiratory Pressure. MEP=Maximum Expiratory Pressure. GAD7=Generalised Anxiety Disorder 7-item scale.

Overall, 45% (323/714) of participants completed both subjective and device-based assessments of sleep quality. When both subjective and device-based groups were compared to each other, and to the broader cohort of participants who consented to research, significant but clinically small differences were observed for age, BMI, deprivation, ethnicity, and COVID-19 severity (**Suppl. Table 1**).

**Table 1:** Cohort demographics for Pittsburgh Sleep Quality Index participants: Participants were categorised by PSQI sleep quality. Continuous values are presented as mean (SD) and were compared using a Wilcoxon rank-sum test. Categorical data are presented as % (n/N) and were compared using a Pearson Chi-squared test. PSQI=Pittsburgh sleep quality index. BMI=body mass index. IMD=Index of multiple deprivation. COPD=Chronic obstructive pulmonary disease. WHO=World health organisation. GAD7=Generalised Anxiety Disorder 7-item scale. Significant *p*-values are shown in bold.

|  |  | **N** | **Good sleep**<br>**quality, N = 258** | **Poor sleep**<br>**quality, N = 456** | **p-**<br>**value** |
| --- | --- | --- | --- | --- | --- |
| PSQI score |  | 714 | 3.4 (1.4) | 10.3 (3.4) | .. |
| Age (years) |  | 704 | 59.6 (13.7) | 57.7 (12.5) | **0.024** |
| Sex (% male) |  | 658 | 70% (166/237) | 55% (230/421) | **<0.001** |
| BMI (kg/m<sup>2</sup>) |  | 632 | 30.7 (6.6) | 32.7 (6.9) | **<0.001** |
| Ethnicity |  | 693 |  |  | 0.74 |
|  | *White* |  | 70% (174/250) | 73% (322/443) |  |
|  | *South Asian* |  | 19% (47/250) | 15% (67/443) |  |
|  | *Black* |  | 6.0% (15/250) | 6.5% (29/443) |  |
|  | *Mixed* |  | 2.8% (7/250) | 2.3% (10/443) |  |
|  | *Other* |  | 2.8% (7/250) | 3.4% (15/443) |  |
| Townsend IMD quintile |  | 703 |  |  | 0.30 |
|  | *1 - most*<br>*deprived* |  | 17% (44/254) | 20% (92/449) |  |
|  | *2* |  | 19% (49//254) | 20% (90/449) |  |
|  | *3* |  | 16% (41/254) | 19% (84/449) |  |
|  | *4* |  | 22% (55/254) | 22% (98/449) |  |
|  | *5 - least*<br>*deprived* |  | 26% (65/254) | 19% (85/449) |  |
| Smoking Status |  | 704 |  |  | 0.82 |
|  | *Never* |  | 60% (153/255) | 58% (260/449) |  |
|  | *Ex-smoker* |  | 39% (99/255) | 41% (182/449) |  |
|  | *Current smoker* |  | 1.2% (3/255) | 1.6% (7/449) |  |
| Average units of alcohol (per week) |  | 710 | 5.9 (7.5) | 4.1 (7.2) | **<0.001** |
| Days since discharge to assessment |  | 714 | 162.0 (38.0) | 162.0 (40.4) | 0.70 |
| **Comorbidities** |  |  |  |  |  |
| Hypertension |  | 650 | 33% (78/237) | 41% (168/413) | 0.060 |
| Diabetes |  | 645 | 18% (43/236) | 24% (98/409) | 0.11 |
| Liver disease |  | 644 | 3.4% (8/236) | 2.9% (12/408) | 0.94 |
| Asthma |  | 648 | 15% (35/236) | 18% (73/412) | 0.40 |
| COPD |  | 647 | 4.2% (10/236) | 4.4% (18/411) | 1.0 |
| Chronic kidney disease |  | 646 | 3.8% (9/237) | 4.4% (18/409) | 0.87 |
| High cholesterol |  | 646 | 24% (56/237) | 24% (97/409) | 1.0 |
| Depression or anxiety |  | 646 | 6.3% (15/237) | 18% (97/409) | **<0.001** |
| **COVID-19 severity** |  |  |  |  |  |
| WHO clinical progression |  | 701 |  |  | 0.52 |
|  | *WHO – class 3-4* |  | 19% (49/255) | 22% (98/446) |  |
|  | *WHO – class 5* |  | 45% (116/255) | 40% (180/446) |  |
|  | *WHO – class 6* |  | 18% (45/255) | 17% (77/446) |  |
|  | *WHO – class 7-9* |  | 18% (45/255) | 20% (91/446) |  |
| Length of stay (days) |  | 711 | 13.5 (16.5) | 15.3 (21.8) | 0.97 |
| ITU admission (% admitted) |  | 706 | 31% (80/257) | 33% (146/449) | 0.77 |
| **Pre-COVID-19 symptoms** |  |  |  |  |  |
| Subjective sleep quality (10=best) |  | 714 | 9.1 (1.8) | 7.4 (2.7) | **<0.001** |
| Subjective dyspnoea (0=best) |  | 712 | 0.9 (1.8) | 1.4 (2.2) | **<0.001** |
| **Post-COVID-19 symptoms** |  |  |  |  |  |
| Subjective sleep quality (10=best) |  | 714 | 8.1 (2.5) | 5.1 (2.9) | **<0.001** |
| Subjective dyspnoea (0=best) |  | 711 | 3.3 (2.8) | 4.5 (2.8) | **<0.001** |
| GAD7 level |  | 696 |  |  | **<0.001** |
|  | *Minimal* |  | 79% (200/252) | 47% (210/444) |  |
|  | *Mild* |  | 17% (42/252) | 23% (104/444) |  |
|  | *Moderate* |  | 3.2% (8/252) | 17% (76/444) |  |
|  | *Severe* |  | 0.8% (2/252) | 12% (54/444) |  |
| Subjective sleep period duration (hours) |  | 706 | 7.4 (1.6) | 6.1 (1.9) | **<0.001** |

Compared to participants reporting good subjective sleep, those with poor sleep quality (PSQI) were more likely to be female, younger, have a higher BMI, previous depression/anxiety, previous dyspnoea, and lower alcohol consumption (**Table 1**).

### Prevalence of Sleep disturbance following COVID-19 hospitalisation

Subjective assessment of sleep quality revealed that 64.0% (456/714) of participants reported poor sleep quality (PSQI). Analysis of temporal changes in sleep quality by the NRS revealed that sleep quality deteriorated following hospital admission for COVID-19 in 53.2% (380/714) of participants. At the early time point sleep quality fell by a median of 3 points and at the late time point sleep quality fell by a median of 2 points (**Fig. 2A**).

**Figure 2:**
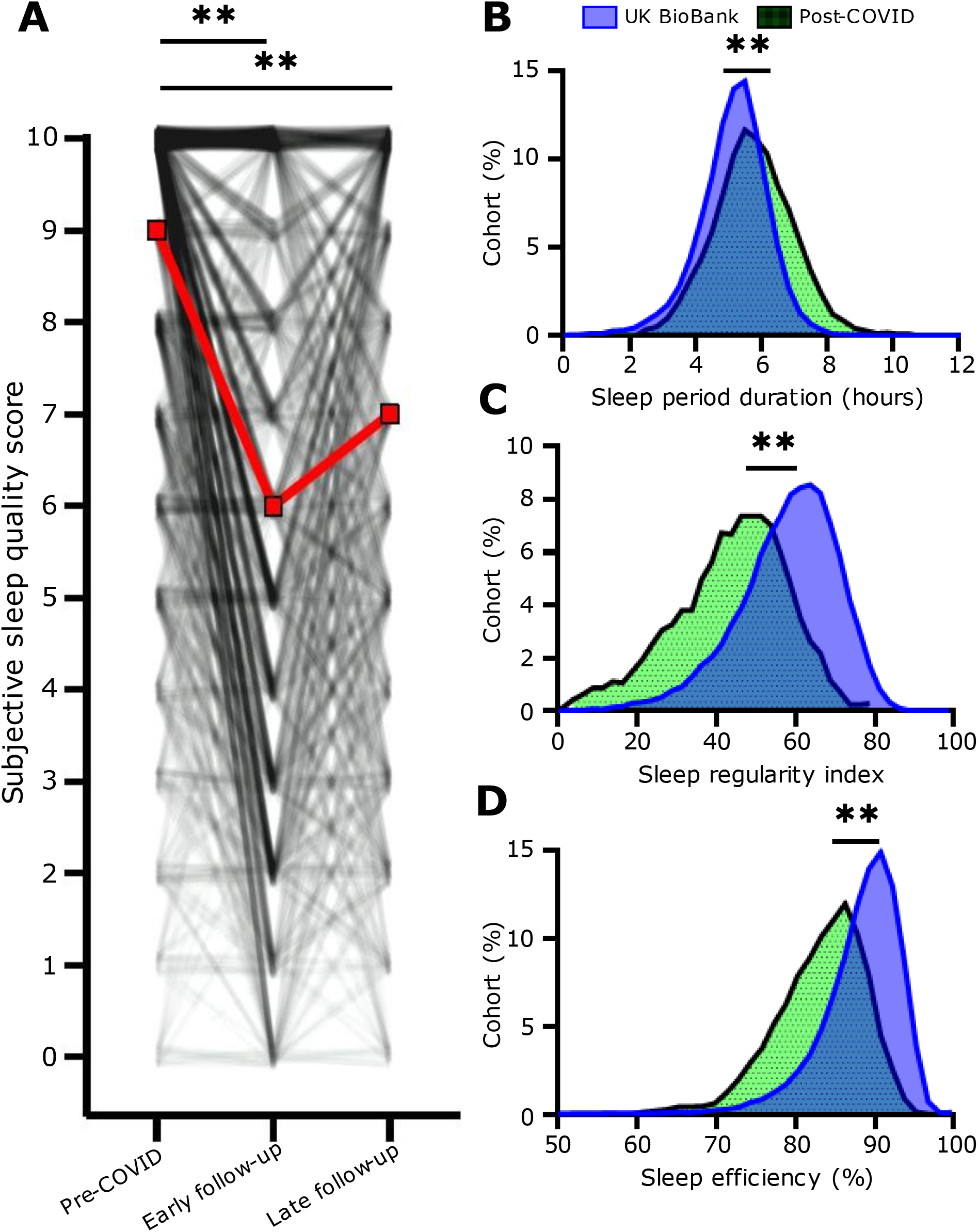
Sleep disturbance after COVID-19 hospitalisation: **(A)** Participants were asked to rate their sleep quality using a numerical rating scale (NRS) either at an early follow-up (median 5 months post COVID-19 for both before COVID and at this time point) as well as at late follow-up (median 12 months post COVID-19). The red line indicates median change, the black lines show individual subjects. **= p<0.001 Dunn’s post-hoc test, Benjamini-Hochberg corrected *p-*-value. Sleep was also quantified using a device-based approach. This was used to quantify **(B)** sleep period duration, **(C)** sleep regularity index and **(D)** sleep efficiency. Our post-COVID cohort is shown in Green with an age-, sex-, and BMI-matched pre-pandemic cohort from the UK BioBank shown in blue (**=p<0.001, t-test).

The actigraphy traces of this cohort were then compared to a UK Biobank cohort, matched for age and sex (demographics in **Suppl. Table 2**). Participants in this study, hospitalised for COVID-19, slept on average 56 minutes longer (**Fig. 2B**), had a lower (−25%) sleep regularity index (**Fig. 2C**) and a lower (3.3 percentage points) sleep efficiency (**Fig. 2D**).

### Relationship of sleep disturbance with dyspnoea

Participants with poor sleep quality (PSQI), scored 4.2 (95%CI 3.0 to 5.5) points (**Fig. 3A)** higher on the dyspnoea-12 questionnaire compared to those with good sleep quality. Sleep deterioration (NRS) was also associated with dyspnoea. Those reporting a deterioration in their sleep quality scored 3.2 (95%CI 2.0 to 4.5) points higher on the dyspnoea-12 questionnaire (**Fig. 3A**) compared to those who did not experience a deterioration. Associations were consistent following adjustments for a minimum set of covariates (age, sex, body mass index (BMI), comorbidities, COVID-19 severity, and length of stay).

**Figure 3:**
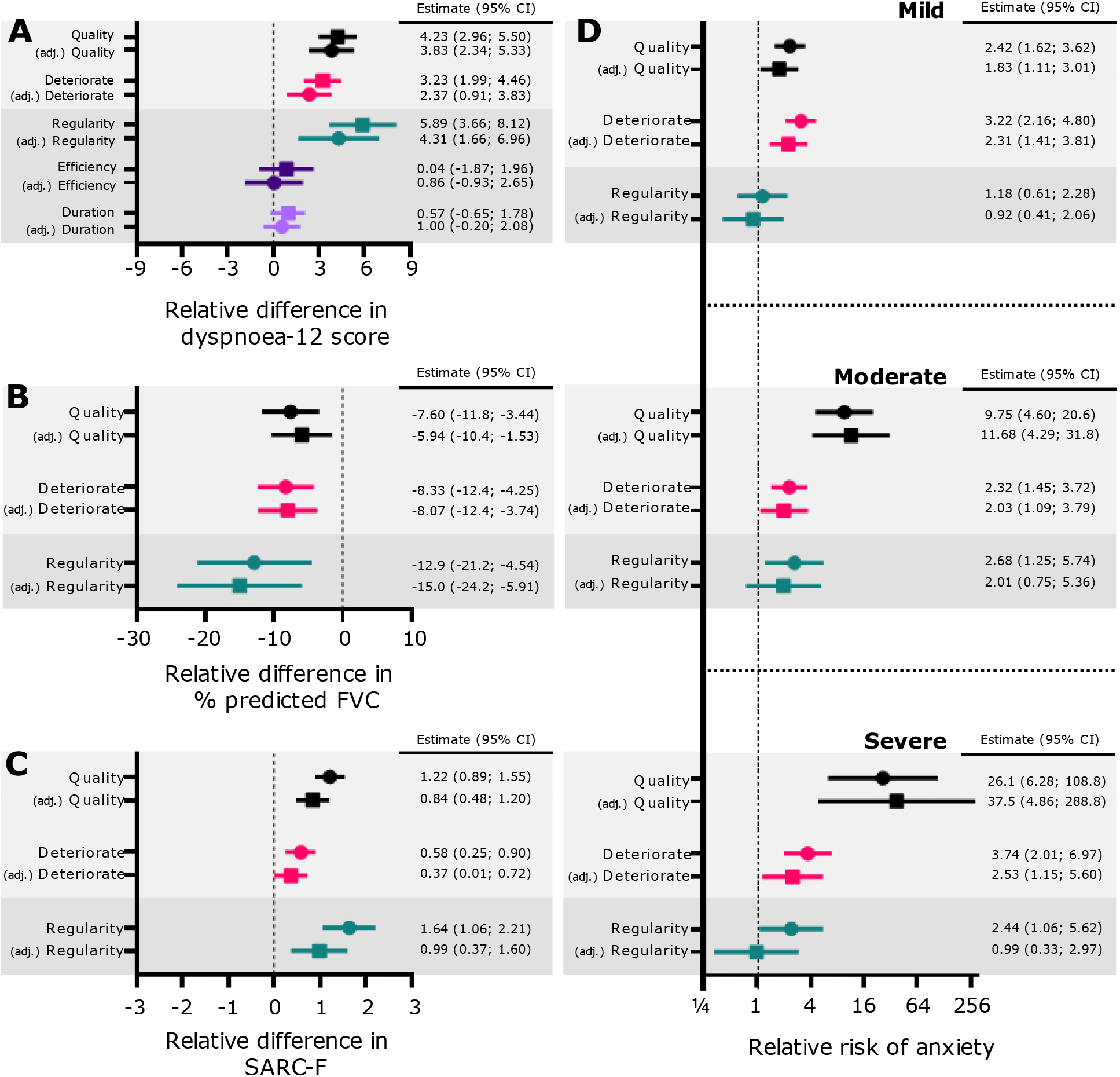
Clinical associations with sleep disturbance: The associations between changes in sleep parameters Sleep quality (PSQI, black); Sleep deterioration (NRS, Pink); Sleep regularity (Teal); Sleep efficiency (Purple); Sleep period duration (Lilac) were investigated for various clinical characteristics. **(A)** Shows the association with Dyspnoea-12 score. **(B)** Shows the association with predicted forced vital capacity (FVC) **(C)** Shows the association with SARC-F score. **(D)** Shows the association with anxiety (GAD-7 scale). Both unadjusted (circles) or multivariable (squares) linear regression (or multinomial logistic regression for anxiety) coefficients are shown. In multivariable linear regression, the association was adjusted for age, sex, BMI, comorbidities, COVID-19 severity, and length of stay. Light grey background indicates subjective evaluation of sleep quality, and a dark-grey background indicates device-based measurement of sleep. BMI=Body Mass Index. FVC=Forced Vital Capacity.

Device-based measurements of sleep were then assessed; participants with the lowest sleep regularity scored 5.9 (95%CI 3.7 to 8.1) points higher on the dyspnoea-12 score compared to participants with the greatest sleep regularity (**Fig. 3A, Suppl. Table 3**). This association was unaffected following adjustment for a minimum set of covariates. No association was observed between dyspnoea and either sleep efficiency or sleep duration in both unadjusted and adjusted models (**Fig. 3A**). Therefore, these measures were not investigated further.

### Relationship of sleep disturbance with lower lung function (FEV^1^ and FVC)

Individuals with poor quality sleep (PSQI) had a lower predicted forced expiratory volume in one second (FEV^1^) of -4.7% (95%CI -9.1 to -0.3%, **Suppl. Fig. 1A**) and a lower predicted forced vital capacity (FVC) of -7.6% (95%CI -11.8 to -3.4%, **Fig. 3B**) compared to those who reported good quality sleep. The association with FEV^1^ was lost following adjustment for a minimum set of covariates (**Suppl. Fig. 1A, Suppl. Table 3**), however, the association with FVC remained (**Fig. 3B, Suppl. Table 3**). Participants who experienced a deterioration in their sleep quality (NRS) following COVID-19 hospitalisation had a lower percent predicted FEV^1^ (−7.9%, 95%CI -12.2% to -3.6%) and a lower percent predicted FVC (−8.3%, 95%CI -12.4% to -4.3%) compared to participants whose sleep quality had remained the same or improved. Associations were consistent following adjustments for the minimal set of covariates (**Fig. 3B, Suppl. Fig. 1A, Suppl. Table 3**).

Sleep regularity was then assessed. Participants with the lowest sleep regularity had a lower percent predicted FVC percent predicted (−12.9%; 95%CI -21.2% to -4.5%; **Fig. 3B**) and a lower percent predicted FEV^1^ (−14.6%; 95%CI -24.3% to -4.8% **Suppl. Fig. 1A**) compared to participants with the highest sleep regularity. This association was also consistent following adjustment for a minimal set of covariates (**Fig. 3B, Suppl. Fig. 1A, Suppl. Table 3**).

Participants’ diffusion capacity was also evaluated. No associations were observed between these measures (KCO, DLCO) and either of the three-sleep metrics for both unadjusted and adjusted models (**Suppl. Fig. 1B, C, Suppl. Table 3**).

### Relationship of sleep disturbance with respiratory pressures

When compared to participants with good quality sleep, those reporting poor quality sleep (PSQI) had a lower maximal expiratory mouth pressure (MEP) (difference: -20.0 cmH^2^O (95%CI -38.6 to -1.4, **Suppl. Fig. 2A**)). In contrast, no difference was observed for maximal inspiratory mouth pressure (MIP) (difference: -11.7 cmH^2^O (95%CI -24.8 to 1.4, **Suppl. Fig. 2B**)). These relationships were consistent following adjustment for a minimal set of covariates (**Suppl. Fig. 2B, Suppl. Table 3**). Furthermore, no associations were observed for either MIP or MEP in those reporting a deterioration in sleep quality (NRS) following COVID-19 hospitalisation (**Suppl. Fig. 2A, B, Suppl. Table 3**).

Participants with the lowest sleep regularity had a lower MEP (−30.1 cmH^2^O, 95%CI -56.3 to -4.0; **Suppl. Fig. 2A**) compared to those participants with the highest sleep regularity. No similar association was observed with MIP. The small sample size (n=55) of this cohort precluded adjustment for a minimal set of covariates.

### Relationship of sleep disturbance with muscle function

Participants with poor sleep quality (PSQI) had a higher score on the SARC-F questionnaire (1.2, 95%CI 0.9 to 1.6; **Fig. 3C**) compared to participants with good quality sleep. Those who reported sleep deterioration (NRS) following COVID-19 hospitalisation also reported higher scores on the SARC-F questionnaire (0.6, 95%CI 0.3 to 0.9 **Fig. 3C**) compared to those participants whose sleep had not deteriorated. Associations were consistent following adjustments for a minimal set of covariates (**Fig. 3C, Suppl. Table 3**). This association was also observed for sleep irregularity. Participants with the most irregular sleep had a higher SARC-F (1.6, 95%CI 1.1 to 2.2 **Fig. 3C, Suppl. Table 3**) score compared to participants with the greatest sleep regularity with similar results following adjustment.

### Relationship of sleep disturbance with anxiety

Participants with poor sleep quality (PSQI) were more likely to have *mild* (Relative Risk (RR) 2.4, 95%CI 1.6 to 3.6), *moderate* (RR 9.8, 95%CI 4.6 to 20.6) or *severe* (RR 26.1, 95%CI 6.3 to 108.8) anxiety compared to participants who reported good quality sleep (**Fig. 3D, Suppl. Table 3**).

A similar association was also observed between anxiety and participants who experienced sleep deterioration (NRS) after COVID-19. Participants who experienced sleep deterioration had a higher relative risk of *mild* (RR 3.2, 95%CI 2.2 to 4.8), *moderate* (RR 2.3, 95%CI 1.5 to 3.7) and *severe* (RR 3.7, 95%CI 2.0 to 7.0) anxiety (**Fig. 3D, Suppl. Table 3**) compared to participants who did not experience deterioration in their sleep quality. Results were consistent following adjustment for a minimal set of covariates.

Participants with the lowest sleep regularity were more likely to report moderate and severe anxiety (*moderate* (RR 2.7, 1.3 to 5.7 95%CI), *severe* (RR 2.4, 1.1 to 5.6 95%CI)) anxiety compared to participants with the highest sleep regularity (**Fig. 3D, Suppl. Table 3**). In contrast, there was no association with *mild* (RR 1.2, 0.6 to 2.3 95%CI) anxiety. Adjustment for the minimal sufficient set of covariates did not affect these associations.

### Mediation analysis for the relationship between sleep disturbance and dyspnoea

Anxiety and altered muscle function are recognised causes of dyspnoea. Mediation analysis was performed (**Suppl. Fig. 3**) to investigate their contribution to mediating the effect between sleep and dyspnoea. Anxiety following COVID-19 mediated the effect of poor sleep quality on dyspnoea by 41.6% (95%CI 25.5 to 60.1%) and reduced muscle function had a similar mediation effect (39.5% (95%CI 24.6 to 57.1%); **Figure 4A, Suppl. Table 4**).

**Figure 4:**
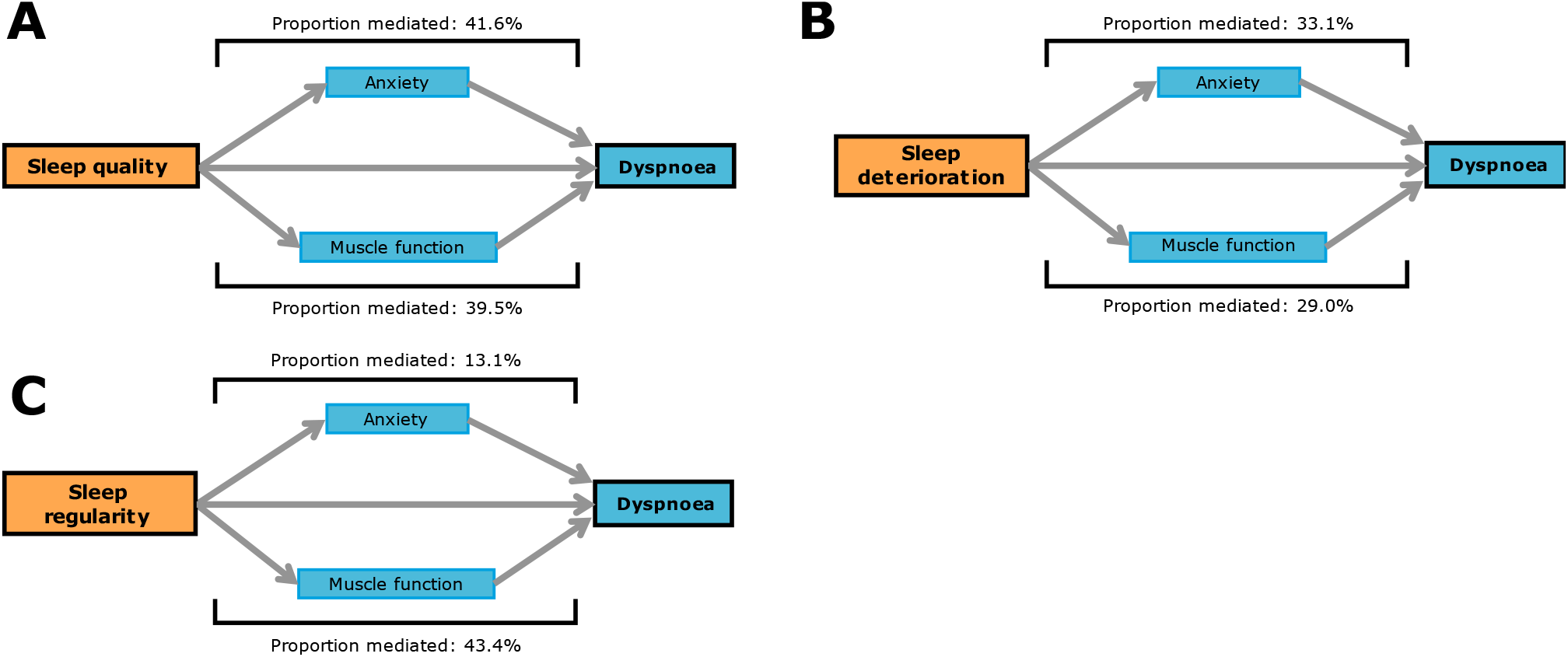
The effect of anxiety or muscle weakness in mediating the effect of sleep on dyspnoea: Mediation models were used to investigate the potential effects of recognised causes of dyspnoea (muscle weakness or anxiety) in mediating the association of sleep on dyspnoea. Three models were performed for the three exposures shown in orange: **(A)** poor sleep quality **(B)** sleep deterioration or **(C)** sleep regularity. Each model had the same outcome, dyspnoea, shown in blue.

For the relationship between sleep quality deterioration and dyspnoea, anxiety mediated the effect by 33.1% (95%CI 23.4 to 43.9%) and reduced muscle function mediated the effect by 29.0% (95%CI 16.7 to 42.3%; **Figure 4B, Suppl. Table 5**). Both anxiety 13.1% (95%CI 1.4 to 29.7%) and reduced muscle function 43.4% (95%CI 20.6 to 70.3%; **Figure 4C, Suppl. Table 6**) also mediated the relationship between sleep irregularity and dyspnoea.

## Discussion

Using multi-modal sleep evaluation conducted in a nationwide UK cohort, we have demonstrated that sleep disturbance is prevalent following hospitalisation for COVID-19. This is likely to persist for at least 12 months as sleep quality did not change between the early (5 months) and late (12 months) follow-up visits. Multi-modal assessment of sleep disturbance revealed that three factors (sleep quality, degradation of sleep quality compared to baseline, and sleep regularity) were associated with dyspnoea and lower lung function. Mediation analysis then revealed that reduced muscle function and anxiety, both recognised causes of dyspnoea^3^, could partially mediate these effects.

Three different complementary methods (PSQI, NRS and device-based)^26^ were used to define sleep disturbance in our study. PSQI is a well-validated assessment tool^33^ that evaluates sleep quality only at the time of administration. Therefore, the evaluation of sleep quality using NRS allowed us to reveal associations in those patients whose sleep quality deteriorated because of hospitalisation complimenting the PSQI analysis. Device-based metrics were then used to investigate specific aspects of sleep quality revealing clinical associations with sleep irregularity. Although the association between dyspnoea and sleep regularity has not been widely reported, sleep regularity outside of the hospitalisation context is known to be associated with pathophysiology^34-36^.

Device-based sleep metrics following hospitalisation for COVID-19 have previously only been measured in participants that had been admitted to critical care^27,28^. Therefore, our cohort now extends these findings revealing altered sleep-based metrics in all participants who had been hospitalised regardless of critical care admission. Both previous studies using device-based metrics in the setting of COVID-19 only revealed clinical associations between anxiety and subjective sleep quality. The lack of association with device-based metrics is an apparent contradiction both with experimental models where sleep disturbance has broad effects^37^ and also clinical studies outside the context of hospitalisation where chronic sleep disturbance is associated with adverse health^38^. Therefore, the discovery of several clinical associations with acute sleep disturbance is in keeping with the wider literature, but also created the possibility that the effect with dyspnoea was mediated through other variables. The mediation analysis suggested that this could only explain part of the effect suggesting that sleep disturbance is directly associated with dyspnoea or mediated via other unidentified clinical or behavioural effects^39^. Further studies will be needed to define this as the association between sleep disturbance and dyspnoea/ lung function has previously but could partially explain the association between sleep disturbance and chronic respiratory disease.

Strengths of our study include its size, multi-centre nature, and the use of different assessment measures to evaluate sleep disturbance. Consistent clinical associations were also observed across each evaluation method therefore the findings cannot be attributed to the use of a specific method. This study does have some limitations which should be considered when interpreting the results. Firstly, the hypothesised directionality of effects in the DAG cannot be confirmed in this study. Although other studies do support these directions^40,41^, bidirectionality of effects have been reported in other settings^16^. NRS quantification of sleep deterioration relied on participant recall and therefore could be affected by recall bias^22^. Selection bias could also affect the results; however, this effect was minimised using bootstrapping combined with cohort matching.

This study provides insight into the prevalence and wider consequences of sleep disturbance following hospitalisation for COVID-19. The findings are likely to translate to other scenarios requiring hospitalisation where sleep disturbance has commonly been reported^15,42,43^, but the effects are poorly understood due to the absence of large cohort studies. The associations described in this study with reduced muscle function, anxiety and dyspnoea suggest that sleep disturbance could be an important driver of the post-COVID-19 condition. If this is the case, then interventions targeting poor sleep quality^44^ could be used to manage multi-morbidity and convalescence following COVID-19 hospitalisation potentially improving patient outcomes.

## Supporting information

PHOSP-COVID collaborative group

## Data Availability

All data produced in the present study are available upon reasonable request to the authors

https://www.phosp.org/

## Author Contributions

The manuscript was initially drafted by CJ, IS, MKR, and JFB, and further developed by the writing committee. CJ, IS, NC, MKR, and JFB made substantial contributions to the conception and design of the work. CJ, IS, RE, JCP, AART, ALH, PLM, RAE, and TP made substantial contributions to the acquisition of data. CJ, IS, TP, PC, ALH, BA-S, RA, CEB, TC, JDC, NC, ABD, GD, CLE, OE, NJG, NAH, VCH, EMH, L-PH, LH-W, LSH, CJJ, MGJ, OCL, KEL, NIL, MM, HJCMc, MAMc, BP, KP-H, KP, BR, MR, PR-O, SR-J, AVR, RMS, JTS, MS, AS, ASh, ASi, SCS, MT, DGW, TY, RGJ, SSi, WD-CM, CEBr, LCW, JCP, AART, AH, PLM, RAE, SEJ, MKR, and JFB made contributions to the analysis or interpretation of data for the work. CJ, IS, RE and TP verified the underlying data. All authors contributed to data interpretation and critical review and revision of the manuscript. All authors had full access to all the data in the study and had final responsibility for the decision to submit for publication.

## Data sharing

The protocol, consent form, definition and derivation of clinical characteristics and outcomes, training materials, regulatory documents, information about requests for data access, and other relevant study materials are available online (https://www.phosp.org/).

## Funding

CJ is funded by an EPSRC DTP (EP/W523884/1). LPH is supported in part by the Oxford NIHR Biomedical Research Centre. AH received support from NIHR Manchester BRC. PLM is supported by an Action for Pulmonary Fibrosis Mike Bray Fellowship. KPH receives funding from Innovate UK (TS/T013028/1) and MRC (MR/W006111/1). JCP receives funding by the NIHR University College London Hospitals Biomedical research centre. BR is funded by the British Heart Foundation Oxford Centre of Research Excellence (RE/18/3/34214). AART is supported by a British Heart Foundation intermediate clinical fellowship (FS/18/13/33281). LVW is supported by GlaxoSmithKline/ Asthma + Lung UK Chair in Respiratory Research (C17-1). DGW is funded by an NIHR Advanced Fellowship (NIHR300669). JFB and PC are supported by an MRC transition support fellowship (MR/T032529/1). JFB is also partially supported by the National Institute of Health Research (NIHR) Manchester Biomedical Research Centre. The work was also supported by an Asthma+Lung UK Malcolm Walleans Grant. The study was also funded by UK Research and Innovation and National Institute of Health Research (grant references: MR/V027859/1 and COV0319).

## Acknowledgements

This study would not be possible without all the participants who have given their time and support. We thank all the participants and their families. We thank the many research administrators, health-care and social-care professionals who contributed to setting up and delivering the study at all of the 65 NHS trusts/Health boards and 25 research institutions across the UK, as well as all the supporting staff at the NIHR Clinical Research Network, Health Research Authority, Research Ethics Committee, Department of Health and Social Care, Public Health Scotland, and Public Health England, and support from the ISARIC Coronavirus Clinical Characterisation Consortium. We thank Kate Holmes at the NIHR Office for Clinical Research Infrastructure (NOCRI) for her support in coordinating the charities group. The PHOSP-COVID industry framework was formed to provide advice and support in commercial discussions, and we thank the Association of the British Pharmaceutical Industry as well NOCRI for coordinating this. We are very grateful to all the charities that have provided insight to the study: Action Pulmonary Fibrosis, Alzheimer’s Research UK, Asthma + Lung UK, British Heart Foundation, Diabetes UK, Cystic Fibrosis Trust, Kidney Research UK, MQ Mental Health, Muscular Dystrophy UK, Stroke Association Blood Cancer UK, McPin Foundations, and Versus Arthritis. We thank the NIHR Leicester Biomedical Research Centre patient and public involvement group and Long Covid Support.

## Notes

### Competing Interest Statement

The authors have declared no competing interest.

### Funding Statement

This study was funded by UK Research and Innovation, National Institute for Health Research, and Engineering and Physical Sciences Research Council.

### Author Declarations

The study was ethically approved by a NHS research ethics committee. The reference is (Ref: 20/YH/0225)

