## Supplementary Methods for "The effects of sleep disturbance on dyspnoea and impaired lung function following COVID-19 hospitalisation: a prospective multi-centre cohort study"

### Supplement

| **Demographics** | | **Overall, N=2,320** | **PSQI+NRS, N=714** | **Device-based, N=829** | ***p*-value[1] (Overall vs PSQ + NRS)** | ***p*-value[2] (Overall vs device-based)** | ***p*-value[3] (PSQ+NRS vs device-based)** |
|---|---|---|---|---|---|---|---|
| Age (years) | | 58.0 (12.6) | 58.4 (13.0) | 59.2 (12.5) | 0.42 | **0.0090** | 0.18 |
| Sex (% male) | | 61% (1,338/2,193) | 60% (396/658) | 63% (497/792) | 0.74 | 0.41 | 0.34 |
| BMI (kg/m2) | | 32.4 (7.3) | 32.0 (6.8) | 31.5 (6.7) | 0.28 | **0.013** | 0.29 |
| Ethnicity | | | | | **0.021** | 0.95 | 0.12 |
| | *White* | 75% (1,685/2,234) | 72% (496/693) | 76% (619/819) | | | |
| | *South Asian* | 12% (262/2,234) | 16% (114/693) | 12% (98/819) | | | |
| | *Black* | 6.9% (154/2,234) | 6.3% (44/693) | 6.1% (50/819) | | | |
| | *Mixed* | 2.1% (46/2,234) | 2.5% (17/693) | 2.2% (18/819) | | | |
| | *Other* | 3.9% (87/2,234) | 3.2% (22/693) | 4.2% (34/819) | | | |
| Townsend IMD quintile | | | | | **0.013** | 0.26 | 0.79 |
| | *1 - most deprived* | 23% (517/2,288) | 19% (136/703) | 21% (177/827) | | | |
| | *2* | 23% (533/2,288) | 20% (139/703) | 21% (171/827) | | | |
| | *3* | 18% (404/2,288) | 18% (125/703) | 17% (143/827) | | | |
| | *4* | 17% (396/2,288) | 22% (153/703) | 20% (164/827) | | | |
| | *5 - least deprived* | 19% (438/2,288) | 21% (150/703) | 21% (172/827) | | | |
| Smoking Status | | | | | 0.17 | 0.86 | 0.13 |
| | *Never* | 55% (1,151/2,105) | 59% (413/704) | 54% (407/759) | | | |
| | *Ex-smoker* | 44% (916/2,105) | 40% (281/704) | 44% (337/759) | | | |
| | *Current smoker* | 1.8% (38/2,105) | 1.4% (38/704) | 2.0% (15/759) | | | |
| Average units of alcohol (per week) | | 4.9 (7.7) | 4.7 (7.4) | 4.7 (7.5) | 0.97 | 0.77 | 0.79 |
| **Comorbidities** | | | | | | | |

| | | | | | | | |
|---|---|---|---|---|---|---|---|
| Hypertension | | 35% (767/2,175) | 38% (246/650) | 35% (275/781) | 0.25 | 1.0 | 0.33 |
| Diabetes | | 22% (473/2,168) | 22% (141/645) | 22% (169/778) | 1.0 | 1.0 | 1.0 |
| Liver disease | | 3.1% (67/2,163) | 3.1% (20/644) | 3.6% (28/778) | 1.0 | 0.58 | 0.72 |
| Asthma | | 19% (408/2,173) | 17% (108/648) | 18% (143/779) | 0.25 | 0.84 | 0.44 |
| COPD | | 5.5% (121/2,171) | 4.3% (28/647) | 5.5% (43/779) | 0.25 | 1.0 | 0.36 |
| Chronic kidney disease | | 4.3% (94/2,172) | 4.2% (27/646) | 4.6% (36/781) | 0.96 | 0.82 | 0.79 |
| High cholesterol | | 20% (442/2,171) | 24% (153/646) | 20% (156/780) | 0.078 | 0.87 | 0.11 |
| Depression or anxiety | | 17% (373/2,168) | 14% (88/646) | 13% (102/778) | **0.036** | **0.0090** | 0.84 |
| **COVID-19 severity** | | | | | | | |
| WHO clinical progression | | | | 0.006 | **<0.001** | 0.059 | |
| | *WHO – class 3-4* | 17% (385/2,273) | 21% (147/701) | 19% (158/820) | | | |
| | *WHO – class 5* | 42% (959/2,273) | 42% (296/701) | 39% (319/820) | | | |
| | *WHO – class 6* | 23% (517/2,273) | 17% (122/701) | 17% (136/820) | | | |
| | *WHO – class 7-9* | 18% (412/2,273) | 19% (136/701) | 25% (207/820) | | | |
| Length of stay (days) | | 13.9 (18.1) | 14.6 (20.1) | 16.4 (21.8) | 0.73 | 0.050 | 0.074 |
| ITU admission (% admitted) | | 33% (701/2,101) | 32% (226/706) | 39% (299/760) | 0.52 | **0.0034** | **0.0038** |
| **Pre-COVID-19 symptoms** | | | | | | | |
| Subjective sleep quality (10=best) | | 7.9 (2.6) | 8.0 (2.5) | 8.1 (2.5) | 0.45 | 0.15 | 0.59 |
| Subjective dyspnoea (0=best) | | 1.2 (2.1) | 1.2 (2.1) | 1.2 (2.1) | 0.69 | 0.56 | 0.89 |
| **Post-COVID-19 symptoms** | | | | | | | |
| Subjective sleep quality (10=best) | | 6.0 (3.1) | 6.2 (3.1) | 6.0 (3.1) | 0.19 | 0.86 | 0.35 |
| Subjective dyspnoea (0=best) | | 4.0 (2.9) | 4.1 (2.8) | 4.1 (2.9) | 0.52 | 0.52 | 1.0 |
| GAD7 level | | | | | 0.43 | 0.26 | 0.66 |
| | *Minimal* | 59% (1,209/2,162) | 59% (410/696) | 60% (578/800) | | | |

| | | | | | | | |
|---|---|---|---|---|---|---|---|
| | *Mild* | 22% (466/2,162) | 21% (146/696) | 19% (150/800) | | | |
| | *Moderate* | 13% (276/2,162) | 12% (84/696) | 12% (98/800) | | | |
| | *Severe* | 9.8% (211/2,162) | 8.0% (56/696) | 9.2% (74/800) | | | |
| Subjective sleep length (hours) | | 6.7 (1.9) | 6.6 (1.9) | 6.6 (1.9) | 0.47 | 0.74 | 0.75 |

**Suppl. Table 1 Demographics of study participants:** The demographics for three groups of participants are shown. The first group, termed Overall, are all participants who consented to research following COVID-19 hospitalisation. The second group, termed subjective, are participants who responded to both the Pittsburgh Sleep Quality Index and Numerical Rating Scale questionnaires. The third group, termed device-based, are participants who had their sleep assessed using actigraphy. Continuous values are presented as mean (±SD) and compared using a Wilcoxon rank-sum test. Categorical data are presented as % (n/N) and compared using a Pearson Chi-squared test. If the value for a patient was missing, that patient was excluded from each specific comparison. PSQI=Pittsburgh sleep quality index. BMI=body mass index. IMD=Index of multiple deprivation. COPD=Chronic obstructive pulmonary disease. WHO=World health organisation. ITU=intensive therapy unit. GAD7=Generalised Anxiety Disorder 7-item scale. Significant *p*-values are shown in bold.

| | | UK Biobank, N=3,080 | Device-based, N=616 |
|---|---|---|---|
| Age (years) | | 65.1 (7.8) | 63.8 (8.5) |
| Sex (% male) | | 65% (2,010/3,080) | 65% (402/616) |
| BMI (kg/m2) | | 30.7 (6.0) | 31.2 (6.3) |
| Ethnicity | | | |
| | *White* | 95% (2,926/3,080) | 77% (474/612) |
| | *South Asian* | 1.5% (45/3,080) | 10% (63/612) |
| | *Black* | 1.8% (55/3,080) | 5.9% (36/612) |
| | *Mixed* | 0.6% (17/3,080) | 2.3% (14//612) |
| | *Other* | 1.2% (37/3,080) | 4.1% (25/612) |
| Smoker (%) | | | |
| | *Never* | 57% (1,742/3,080) | 52% (299/576) |
| | *Ex-smoker* | 35% (1,074/3,080) | 47% (268/576) |
| | *Current smoker* | 8.3% (257/3,080) | 1.6% (9/576) |
| Subjective sleep length (hours) | | 7.1 (1.1) | 6.6 (1.8) |
| **Comorbidities** | | | |
| Hypertension | | 24.5% (754/3,080) | 41% (246/607) |
| Diabetes | | 5.1% (158/3,080) | 23% (137/605) |
| Liver disease | | 0.3% (8/3,080) | 3.8% (23/605) |
| Asthma | | 13.9% (427/3,080) | 17% (105/605) |
| COPD | | 1% (32/3,080) | 6.8% (41/605) |
| Chronic kidney disease | | 0.1% (4/3,080) | 4.8% (29/606) |
| High cholesterol | | 9.2% (284/3,080) | 22% (136/606) |
| Depression or anxiety | | 6.4% (197/3,080) | 12% (75/606) |

**Suppl. Table 2 Demographics of participants compared to the UK biobank:** The device-based cohort were compared to the UK Biobank control group which was matched for age, sex, and BMI. Note that the matching was done at an aggregated level. Continuous values are presented as mean (SD). Categorical data are presented as % (n/N). If the value for a patient was missing, that patient was excluded from each specific comparison. BMI=Body Mass Index. COPD=Chronic obstructive pulmonary disease.

| | Sleep quality (PSQI) | Sleep deterioration (NRS) | Sleep regularity |
|---|---|---|---|
| Dyspnoea-12 | 3.05 (2.36; 3.75) | 1.49 (0.77; 2.21) | 1.32 (0.66; 1.99) |
| Predicted $FEV_1$ (%) | -2.41 (-4.75; -0.07) | -4.05 (-6.28; -1.82) | -4.70 (-6.86; -2.55) |
| Predicted FVC (%) | -3.65 (-5.84; -1.47) | -3.95 (-6.06; -1.85) | -4.64 (-6.80; -2.48) |
| Predicted TLCO (%) | -0.33 (-7.11; 6.45) | -1.74 (-5.84; 2.35) | 0.02 (-3.79; 3.83) |
| Predicted KCO (%) | -0.85 (-5.31; 3.62) | 0.20 (-6.05; 6.45) | 2.95 (-2.98; 8.88) |
| MIP | -9.14 (-16.8; -1.49) | -3.35 (-11.2; 4.49) | .. |
| MEP | -12.8 (-23.8; -1.89) | 1.21 (-10.2; 12.6) | .. |
| SARC-F | 0.67 (0.50; 0.84) | 0.21 (0.04; 0.39) | 0.39 (0.22; 0.56) |
| GAD7 (Mild) | 1.53 (1.16; 2.01) | 1.79 (1.11; 2.89) | 0.92 (0.41; 2.06) |
| GAD7 (Moderate) | 3.91 (2.69; 5.68) | 3.62 (1.95; 6.73) | 2.01 (0.75; 5.36) |
| GAD7 (Severe) | 7.54 (4.63; 12.3) | 4.90 (2.26; 10.6) | 0.99 (0.33; 2.97) |

**Suppl. Table 3 Dyspnoea, muscle weakness and anxiety are associated with sleep disturbance:** In contrast to the rest of the paper the evaluations (PSQI, NRS and device-based) for sleep disturbance were treated as a continuous variable**.** Multivariable linear regression was used to evaluate the standardised effect estimates for all variables except anxiety (GAD7); multinomial logistic regression was used to evaluate the relative risk with anxiety. In all cases, the estimates were standardised by centring and scaling (subtract mean, divide by standard deviation) the sleep variable and all models were adjusted for a minimally sufficient set of covariates. $FEV_1$=Forced Exhaled Volume in 1 second. FVC=Forced Vital Capacity. MIP=Maximum Inspiratory Pressure. MEP=Maximum Expiratory Pressure. GAD7=Generalised Anxiety Disorder 7-item scale. Missing results are due to the sample size being too small to adjust for a minimally sufficient set of covariates.

| Label | Effect size | Mediated effect | Proportion mediated, % |
|---|---|---|---|
| $c'$ | 0.9 (-0.3; 2.0) | | |
| $a_1$ | 0.7 (0.6; 0.9) | 1.9 (1.2; 2.8) | 41.6 (25.5; 60.1) |
| $b_1$ | 2.7 (1.1; 1.9) | | |
| $a_2$ | 1.2 (0.9; 1.6) | 1.8 (1.1; 2.6) | 39.5 (24.6; 57.1) |
| $b_2$ | 1.5 (1.1; 1.9) | | |

**Suppl. Table 4:** Results of the multiple mediation model using poor sleep quality (PSQI) as the exposure and dyspnoea as the outcome. Results show 95% confidence intervals calculated using a bootstrap with 1,999 resamples.

| Label | Effect size | Mediated effect | Proportion mediated, % |
|---|---|---|---|
| $c'$ | 1.2 (0.5; 1.8) | | |
| $a_1$ | 0.4 (0.3; 0.5) | 1.0 (0.7; 1.3) | 33.1 (23.4; 43.9) |
| $b_1$ | 2.3 (1.8; 2.7) | | |
| $a_2$ | 0.6 (0.3; 0.8) | 0.9 (0.5; 1.3) | 29.0 (16.7; 42.3) |
| $b_2$ | 1.6 (1.4; 1.8) | | |

**Suppl. Table 5:** Results of the multiple mediation model using sleep deterioration (NRS) as the exposure and dyspnoea as the outcome. Results show 95% confidence intervals calculated using a bootstrap with 1,999 resamples.

| Label | Effect size | Mediated effect | Proportion mediated, % |
|---|---|---|---|
| $c'$ | 2.8 (0.4; 5.1) | | |
| $a_1$ | 0.4 (0.1; 0.6) | 0.8 (0.1; 1.9) | 13.1 (1.4; 29.7) |
| $b_1$ | 2.3 (0.8; 4.0) | | |
| $a_2$ | 1.5 (0.9; 2.1) | 2.8 (1.3; 4.5) | 43.4 (20.6; 70.3) |
| $b_2$ | 1.9 (1.2; 2.5) | | |

**Suppl. Table 6:** Results of the multiple mediation model using sleep regularity as the exposure and dyspnoea as the outcome. Results show 95% confidence intervals calculated using a bootstrap with 1,999 resamples.

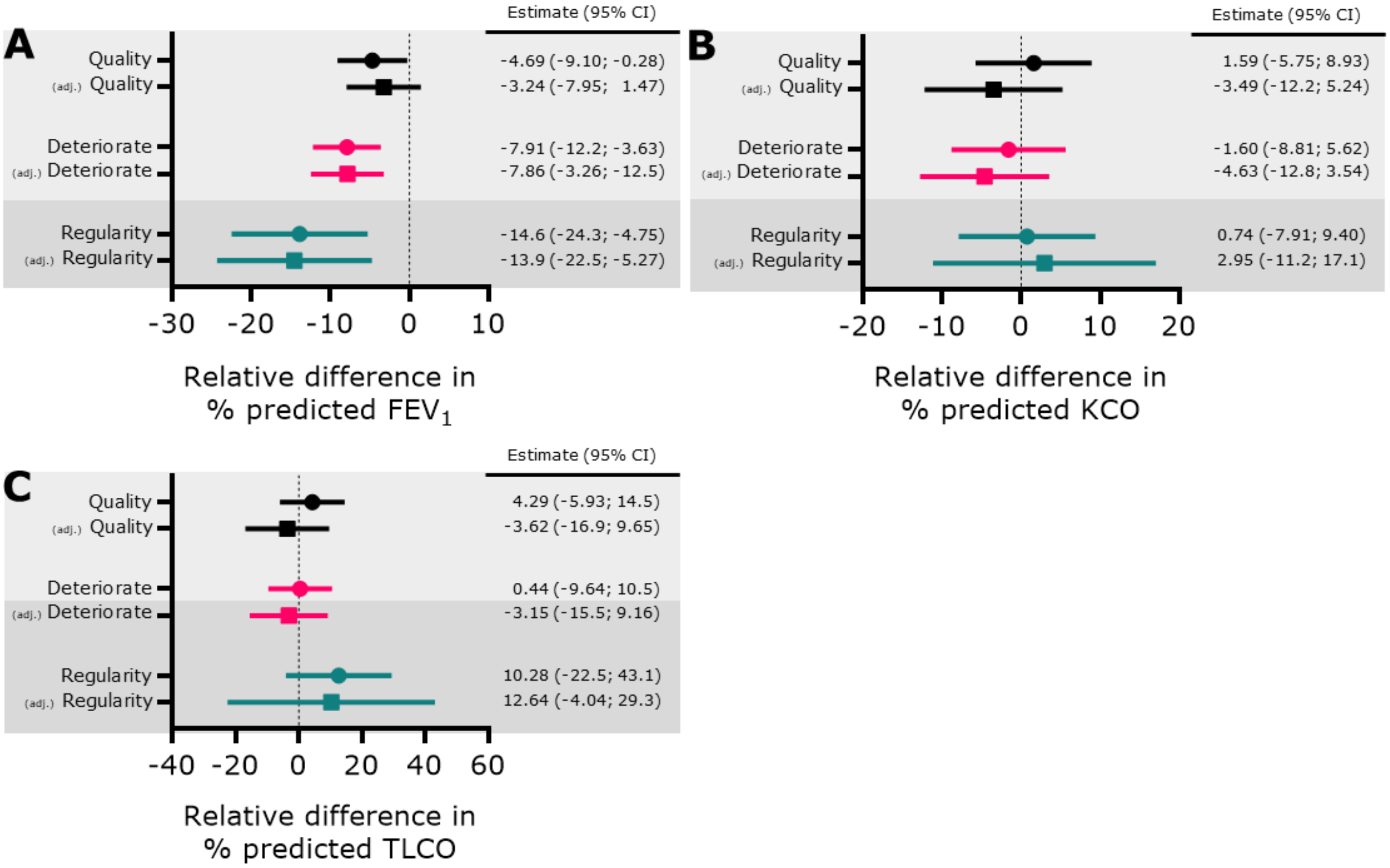


**Supplementary Figure 1: Sleep disturbance is associated with altered lung function:** The associations between changes in the sleep parameters Sleep quality (PSQI, black); Sleep deterioration (NRS, Pink); Sleep regularity (Teal) were investigated between sleep disruption and lung function **(A)** Shows the association with percentage predicted $FEV_1$. (**B)** Shows the association with KCO **(C)** Shows the association with TLCO. Both unadjusted (circles) or multivariable (squares) linear regression coefficients are shown. In multivariable linear regression, the association was adjusted for age, sex, BMI, comorbidities, COVID-19 severity, and length of stay. Light grey background indicates subjective evaluation of sleep quality, and a dark-grey background indicates device-based measurement of sleep. BMI=Body Mass Index. $FEV_1$= Forced Expiratory Volume in 1 second, KCO= Carbon monoxide transfer coefficient, TLCO= Transfer capacity of the lung.

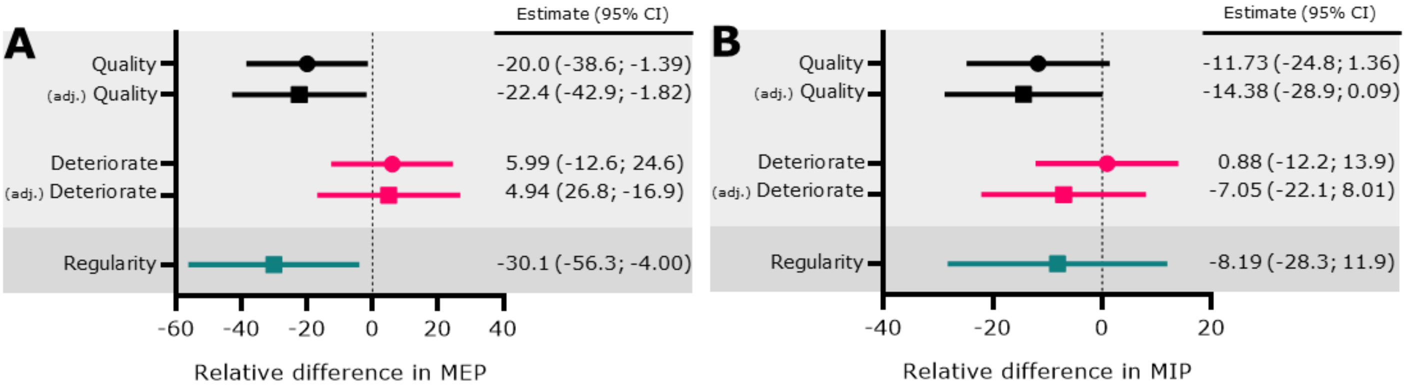


**Supplementary Figure 2: Sleep disturbance is associated with altered respiratory pressures:** Associations between changes in the sleep parameters Sleep quality (PSQI, black); Sleep deterioration (NRS, Pink); Sleep regularity (Teal) were investigated with altered respiratory pressures. **(A)** Shows the association with maximal expiratory pressure (MEP). **(B)** Shows the association with maximal inspiratory pressure (MIP). Both unadjusted (circles) or multivariable (squares) linear regression coefficients are shown. In multivariable linear regression, the association was adjusted for age, sex, BMI, comorbidities, COVID-19 severity, and length of stay. A light grey background indicates the variable is a subjective measure and a dark-grey background indicates a device-based measure.

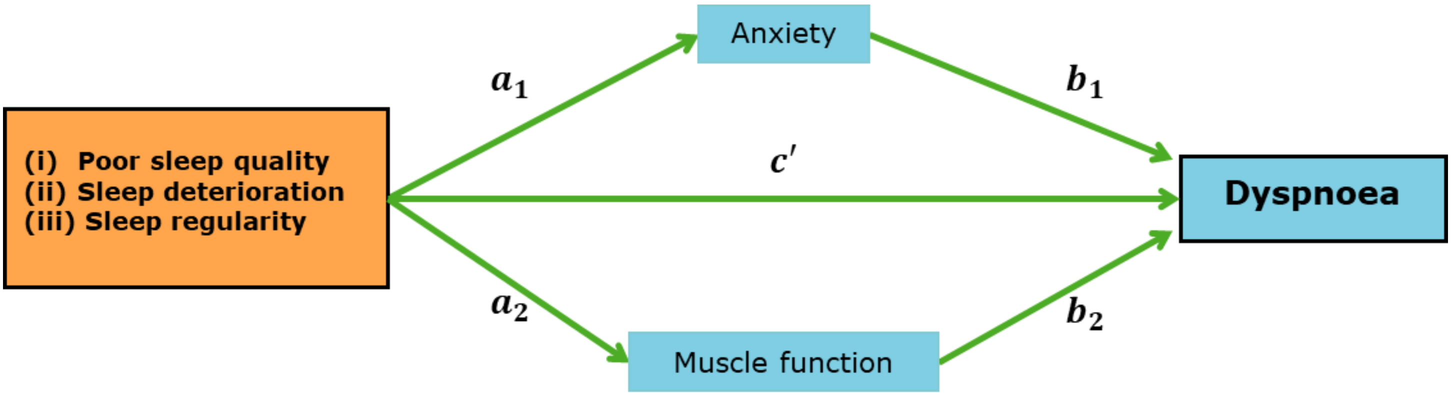


**Supplementary Figure 3: Mediation model:** Mediation models investigated the association between sleep disruption and dyspnoea to investigate whether anxiety and muscle function could be considered mediators in the relationship. The results are reported in **Suppl. Tables 4-6** as well as in **Figure 4.**

**Supplementary Methods:**

**Participants:** Participants who consented to research were enrolled in the study. For these participants, follow-up data were collected at two time points: an early time point 2-7 months after hospital discharge, and a later time point 10-14 months after hospital discharge. According to the site where a participant was enrolled, they could also complete questionnaires, such as the Pittsburgh Sleep Quality Index (PSQI) questionnaire or participate in actigraphy data collection. Sex was classified as sex assigned at birth. Age was categorised as < 40, 40–49, 50–59, 60–69, and ≥70 years; body mass index (BMI) was categorised as *underweight* (< 20 kg/m$^2$), *normal weight* (20-24.9 kg/m$^2$), *overweight* (25-29.9 kg/m$^2$), *obese* (30-34.9 kg/m$^2$) and *morbidly obese* (>34.9 kg/m$^2$); ethnicity was recorded according to census definitions and categorised as the four most frequent classes in the dataset: *White*, *South Asian*, *Black*, *Mixed* and *Other*; the Townsend index of multiple deprivation (IMD) was categorised as quintiles with the first quintile indicating the most deprived; length of hospital stay was categorised as quartiles with the first quartile indicating the shortest stays; five comorbidities were analysed as binary *Yes* or *No* variables, based on the medical history of the participants.

**Subjective assessment of sleep quality:**

**(ii) Numerical rating scale:** Participants answered the question "*For each symptom, please rate them before you had COVID-19 and how you are now*". At the early assessment time point, participants rated their sleep quality and recalled their pre-COVID-19 sleep quality. At the late assessment time point, participants rated their current sleep quality.

**Actigraphy:** Tri-axial acceleration data were collected at a 30Hz sampling frequency and were processed using the GGIR package (version 2.2-0, http://cran.r-project.org) in R[1-3]. The GGIR package is a sleep detection algorithm validated with polysomnography (PSQ)[2] and has automated detection of the sleep period time window[3]. The R script used to produce estimates of sleep regularity index, sleep efficiency and sleep period duration has previously been described and is open source[4]. The definitions of these three variables are:

| **Sleep metric** | **Definition** |
|---|---|
| **Sleep regularity index[5]** | The sleep regularity index is the percentage probability that a participant is in the same state (asleep or awake) at any two time points exactly 24 hours apart. |

| | |
|---|---|
| | A participant who is asleep/awake at the exact same times every day would score 100. |
| **Sleep period duration** | Sleep period duration is the length of the sleep period time window. That is the time between the onset of sleep and the final wake-up of the night. |
| **Sleep efficiency** | Sleep efficiency is the percentage of time in the sleep period duration that a participant was estimated to be asleep.<br><br>A participant who had no night-time arousals (e.g., never rolling over) would have 100% efficiency. |

The same settings for the algorithms were used for the analysis in PHOSP and the UK Biobank cohort[6].

**Symptom assessment:**

| Symptom | Assessment | Interpretation |
|---|---|---|
| **Dyspnoea** | **Dyspnoea-12 questionnaire**<br><br>Dyspnoea was assessed using the dyspnoea-12 questionnaire[7], incorporating both physical and affective aspects, consisting of 12 descriptor questions, each scored 0 – 3. | A higher score indicates worse dyspnoea |
| **Lung Function** | Performed according to local standard operating procedures dependent on the availability of local equipment. The highest reading recorded per participant was used in this study and percent predicted values were calculated using the Global Lung Function Initiative (GLI) reference equations[8] | |
| **Anxiety** | **Generalised Anxiety Disorder 7-item assessment**[9]<br><br>Anxiety was assessed using the generalised anxiety disorder 7-item assessment which has a scale ranging from 0 – 21. | The results were categorised:<br>Minimal (0-4)<br>Mild (5-9)<br>Moderate (10-14)<br>Severe (≥15)[9] |
| **Muscle Function** | **SARC-F questionnaire**<br>This was performed as previously described[10]. | A lower score indicates better muscle function. |

**UK Biobank cohort:** Actigraphy was performed in the UK biobank between February 2013 and December 2015 when all participants with a valid e-mail address were invited to wear a wrist-worn accelerometer device (Axivity AX3) on their dominant wrist 24/day for seven days. Tri-axial acceleration data were collected at 100Hz Of the total cohort, 20.6% (103,713/502,540) of participants were accepted. Further details of data collection and processing have been previously reported[11]. The traces were analysed using GGIR[1], as described above, using identical parameters to the PHOSP analysis. When comparing the PHOSP-COVID cohort to the pre-pandemic UK Biobank cohort, an age-, sex-, and BMI-matched aggregated cohort was selected from the UK Biobank. This cohort was age-, sex-, and BMI0matched at an aggregate level (using the age and BMI categories described above). Due to the UK Biobank only enrolling patients aged 40 – 69, all PHOSP participants aged <40 could not be matched. A ratio of 1:5 participants (PHOSP-COVID: UK BioBank) was chosen. Both the PHOSP-COVID cohort and UK BioBank cohort were split into sub-cohorts based on each age-, sex-, and BMI-combination (i.e., group 1: 40 – 49, Males, Normal Weight, group 2: 40 – 49, Males, Overweight, …, group 6: 40 – 49, Females, Normal Weight, …). Since the UK BioBank sub-cohorts always contained at least 5 times more participants than the PHOSP sub-cohorts, the required number of participants were selected at random from the UK Biobank to generate the matched cohort. This process was repeated 25 times so that 25 separate matched cohorts could be considered. This was done as a sensitivity analysis to ensure random sampling had not generated an unrepresentative sample. The results of only a single cohort are reported but all analyses gave rise to the same results.

**Actigraphy data collection between PHOSP-COVID and the UK Biobank**

PHOSP-COVID protocol used the GENEActiv Original device, worn on the non-dominant wrist for 14 days. This contrasts with the UK Biobank protocol which used the Axtivity AX3, worn on the dominant wrist for 7 days. Differences between different devices worn on different wrists have previously been explored, and it was concluded that sleep outcomes from this combination of non-dominant wrist GENEActiv and dominant wrist Axtivity AX3 fell within the proposed equivalence (10% equivalency)[12]. The effect of different study lengths on certain sleep metrics has also been investigated. The metrics considered in this paper are *consecutive* measures, that is they capture variability in sleep-wake patterns between consecutive days, which are stable across varying study lengths[13].

**DAG and covariates:**

Potential covariates for associations between sleep disturbances and dyspnoea were selected using a Directed Acyclic Graph (DAG, **Suppl. Material**). This was constructed based on previous literature and consulting subject matter experts. This suggested the minimally sufficient adjustment set

- Age, sex, BMI, deprivation, length of stay, WHO COVID severity, and pre-COVID comorbidities.

The comorbidities that were included are high cholesterol, diabetes, asthma, hypertension, and depression/anxiety. These were derived from the medical history taken during hospital admission and are treated as five binary *Yes/ No* variables.

**Regression bootstrap confidence intervals**

All associations were analysed using linear regression both unadjusted and adjusting for the minimally sufficient set of covariates. Due to the residual's departure from normality (assessed via QQ-plot), bootstrap 95% confidence intervals for the regression coefficients were used. The approach used here was to bootstrap the residuals of the fitted model. If you are seeking to estimate the coefficient $\beta_1$ of the straight line $y = \beta_0 + \beta_1 x$, the method is:

1. Fit the model to the original data $(x_1, y_1), (x_2, y_2), \ldots, (x_n, y_n)$ to get coefficient estimates $\hat{\beta}_0$, $\hat{\beta}_1$, residuals $r_1, r_2, \ldots, r_n$ and the estimate for the error variance $s^2 = (n-2)^{-1} \Sigma_{j=1}^{n} r_j^2$.
2. To generate your bootstrap sample:
   a. Set $x_j^* = x_j$;
   b. Randomly sample $\varepsilon_j^*$ from $\{r_1 - \bar{r}, r_2 - \bar{r}, \ldots, r_n - \bar{r}\} \times s$, where $\bar{r}$ is the mean of the residuals and $s$ is the square root of the estimated error variance.
   c. Set $y_j^* = \hat{\beta}_0 + \hat{\beta}_1 x_j + \varepsilon_j^*$ (i.e., the fitted value from the original model plus a randomly selected $\varepsilon_j^*$).
   d. Repeat steps (a) – (c) until a full sample $(x_1^*, y_1^*), (x_2^*, y_2^*), \ldots, (x_n^*, y_n^*)$ has been generated
3. Calculate the least squares estimate and standard errors, $\hat{\beta}_1^*$ and $SE(\hat{\beta}_1^*)$, respectively, based on the bootstrap sample generated in step (2).
4. Calculate the $Z$-score

$$Z_i^* = \frac{\hat{\beta}_1^* - \hat{\beta}_1}{SE(\hat{\beta}_1^*)}$$

Repeat steps (1) – (3) to generate the desired number of bootstrap estimates (here, $B = 1999$ bootstrap samples were calculated). This will result in the sample $Z_1^*, Z_2^*, \ldots, Z_B^*$. To calculate the $100(1-2\alpha)\%$ confidence interval for $\beta_1$, compute the empirical $\alpha$- and $(1-\alpha)$-quantiles of $Z_1^*, Z_2^*, \ldots, Z_B^*$. Due to the choice of $B = 1999$, $\hat{t}^{(\alpha)}$ is the $(1999 + 1) \times 0.025 = 50^{\text{th}}$ smallest $Z_j^*$ and $\hat{t}^{(1-\alpha)}$ is the $(1999 + 1) \times 0.975 = 1950^{\text{th}}$ smallest (i.e., the 50th largest) $Z_j^*$. This gives the $100(1-2\alpha)\%$ confidence interval for $\beta_1$ as:

$$\left(\hat{\beta}_1 - \hat{t}^{(1-\alpha)}\, SE(\hat{\beta}_1),\ \hat{\beta}_1 - \hat{t}^{(\alpha)}\, SE(\hat{\beta}_1)\right).$$

**Mediation analysis**

A multiple mediation model was used to estimate the direct effect of sleep disturbance metrics, and the indirect effects of anxiety and muscle function, on dyspnoea. Three separate exposures were considered: (i) Poor sleep quality (PSQI), (ii) sleep deterioration (NRS), and (iii) sleep regularity. This led to three multiple mediation models (**Suppl. Fig. 4**). Linear regression with the product of coefficients method was used which estimates the effect of the exposure on the mediators ($a_i$; $i = 1,2$), the effect of the mediators on the outcome ($b_i$; $i = 1,2$) and the direct effect of the exposure on the outcome after adjusting for the mediators ($c'$). The average causal mediation effect (ACME) for each mediator is then calculated as $\gamma_i = a_i \times b_i$; $i = 1,2$ and the proportion mediated by the $i^{\text{th}}$ mediator is $100 \times \gamma_i/(c' + \gamma_1 + \gamma_2)$. Confidence intervals for the ACMEs were computed using a bootstrap approach to loosen the assumption of normally distributed residuals with 1,999 resamples. This was conducted using the R package *lavaan* version 0.6-12[14].
